# Identifying missed opportunities in tuberculosis preventive treatment care cascade: Analysis of programme data from Maharashtra, India

**DOI:** 10.1101/2025.04.24.25326379

**Authors:** Anuj Mundra, Tarun Bhatnagar, Mrinalini Das, Sandeep Sangale, Hemant Patil, Abhishek Raut, Aniruddha Kadu, Rameshwar J Paradkar, Subodh S Gupta, Bishan S Garg

## Abstract

Tuberculosis infection is a condition when a person harbours the bacilli without having signs of active disease. In India, approximately 70% of household contacts of people with pulmonary tuberculosis have the infection. The national tuberculosis elimination programme recommends preventive treatment to household contacts of all people with pulmonary tuberculosis after ruling out active disease. Maharashtra is one of the bigger states in India with high tuberculosis burden.

We analysed the state programme data to describe the tuberculosis preventive treatment care cascade for household contacts of all notified people with pulmonary tuberculosis for the year 2023 in Maharashtra.

Contact tracing was done for 84% of the 1,33,167 notified people with pulmonary tuberculosis. A total of 4,06,291 household contacts were enlisted out of whom 3,86,224 (95%) were screened for symptoms of tuberculosis. 185502 (45%) household contacts were listed as eligible for tuberculosis preventive treatment, of whom 101325 (55%) were initiated on tuberculosis preventive treatment. While 41,480 (41%) of those initiated on treatment successfully completed it, treatment outcomes were not recorded for around 57,191 (56%) of them. Tuberculosis preventive treatment completion as well as recording of treatment outcomes was lesser for 6H regimen, among contacts of those seeking care from private sector and clinically diagnosed people with tuberculosis.

Reasons for losses from the cascade need to be identified and addressed. Addressing missing data will further help in understanding programme performance. Frontline workers may be trained and utilized to capture real time data using simpler tools. Aligning the annual India TB report with the guidelines by including household contacts of all notified persons with pulmonary tuberculosis instead of only microbiologically positive ones may improve treatment outcome recording among clinically diagnosed cases. The capacity building, monitoring, and supportive supervision need further strengthening to improve the provision of tuberculosis preventive treatment care.

## Introduction

The Sustainable Development Goals targets to control tuberculosis (TB) as a public health problem and envisages a reduction in the incidence rates by 80% of 2015 levels by the year 2030.[1] Tuberculosis infection (TBI) is a state where a person harbours the causative bacilli, and demonstrates immune response against it, without signs of active disease.[2] An estimated 2 billion people globally are infected with TB.[3] About 10% of those with TBI develop TB disease sometime in near future.[2] Household contacts (HHC) of people with TB are at higher risk of getting infected. Previous studies have estimated that around 14-20% of transmission of TBI could be attributed to household transmission.[4–6] According to a systematic review, the prevalence of TBI among household contacts (HHC) of people with TB globally is 42%.[7]

India aims to achieve the Sustainable Development Goals 2030 TB target by the year 2025.[8,9] This needs an accelerated rate of annual decline in the TB incidence. The National Tuberculosis Elimination Programme (NTEP) is employing multiple strategies to achieve this ambitious target.[9] About 70% of HHC of people with pulmonary TB (PwPTB) have been reported to have TB infection and eventually 12 HHC per 1000 person-years developed the disease.[10] One recent strategy of NTEP is focusing on screening of TB and TPT provision in HHC (aged more than 6 years) of PwPTB, considering they are one of the groups at risk of developing TB. The Guidelines for Programmatic management of TPT was launched in India in the year 2021 to reduce TBI transmission in line with the END TB strategy.[1,9]

As this is a newer initiative, evidences of TBI and TPT management in routine programme settings are few. A recent operational research discusses the risk factors for TPT non completion from West Bengal, India.[11] Some studies describe the losses in TPT care cascade among children and its reasons.[12,13] But studies discussing the same for all HHC are limited to few districts of Maharashtra, Karnataka and Delhi have reported several challenges pertaining to the processes, logistics, training, community engagement and acceptance leading to losses in the care cascade and thus missed opportunities.[14–16]

In India, the state of Maharashtra is one of the high TB burden states contributing to 12% of country’s burden of PwPTB (0.15 million) in 2023. In the same year, the state could put 48000 HHC on TPT, which represents 6% of the HHC initiated on TPT in India.[17] This suggests losses during the TPT care cascade, which needs to be quantified at each step to understand the current TBI and TPT situation in Maharashtra. We believe, evidence of TPT care cascade from a large, high TB burden state will help developing strategies or interventions for TBI management and achieve the desired TB control in India.

We therefore planned this study with the objective of describing the TPT care cascade for HHC of all notified PwPTB for the year 2023 in the state of Maharashtra. Based on the findings of this study, we plan to develop and implement interventions in routine programme settings to strengthen the care cascade.

## Methods

### Study Settings

#### General setting

Maharashtra is one of the bigger state in India with an area of about 3,00,000 square kilometers and a population of 130 million across 36 administrative districts divided administratively into 6 divisions.[18] The state in situated in western part of the country, bordering six other states and the Arabian sea on one side. The state comprises 45% urban population and 9% tribal population.[19]

#### Specific setting

Maharashtra has 80 NTEP districts providing services through 528 TB units. The state follows the national guidelines, according to which, once contact tracing is done, the HHC, who after exclusion of TB disease are found positive for TBI through tuberculin sensitivity test or interferon gamma release assay are eligible for receiving TPT. However, those who are unable to get these tests done are also provided with TPT at the earliest after ruling out active TB disease. In addition to 6H regimen (6 months of daily isoniazid), new shorter regimens of TPT are provided to the eligible HHC. Contact tracing is the primary responsibility of frontline workers viz. ASHA worker (Accredited Social Health Activist), screening of the enlisted contacts and referral for further assessment of eligibility is the primary responsibility of health and wellness centre – sub centre staff including community health officers. Further assessment of the contacts for TBI or TB disease as applicable is to be done at PHC or higher levels. The staff from TB unit including senior treatment supervisor is responsible for capacity building and supportive supervision of field staff and support for treatment adherence.[9]

The senior treatment supervisor uses *Ni-kshay* portal, a web-based patient management system for recording and reporting data related to TPT cascade for HHC in 3 electronic registers.[20] All index PwPTB are registered in TB notification register and identified by a unique patient id. Whether contact tracing was done for the index PwPTB is also recorded here. The contact tracing register records the aggregate number of contacts identified for each PwPTB, those screened for TB, evaluated for TB and TBI, eligible for TPT and numbers initiated on TPT. The TPT register contains the individual details of all persons who are initiated on TPT along with the dates and treatment outcomes.

#### Study design

A retrospective cohort study was conducted based on routinely collected programmatic data extracted from the notification register, contact tracing register, and TPT register of the web-based patient management system of NTEP (*Ni-kshay)*.

#### Study population

The study population included 1) index PwPTB diagnosed and notified in Maharashtra between January and December 2023 and 2) household contacts of all PwPTB notified between January and December 2023.

#### Data management and analysis

Data, extracted in MS excel format, from the three registers were imported in SPSS v20 software. Data from notification and contact tracing registers were merged using the unique patient id. People with extra pulmonary TB, site not recorded and those notified outside the specified period (Jan-Dec 2023) were excluded. All the identifiers other than unique patient id were removed prior to performing the analysis. The data was downloaded in November 2024.

For the purpose of this study, we operationally classified all treatment outcomes other than ‘TPT completed’ (died, lost to follow-up, treatment failed, TPT discontinuation due to toxicity, and not evaluated) as ‘TPT not completed’. Categorical variables were summarized as frequency and percentages. Continuous variables were summarized as median (Inter-quartile range, IQR).

#### Ethics statement

Ethics approval was obtained from Institutional human ethics committee, National Institute of Epidemiology, Chennai (NIE/IHEC/A/202408-03 dated 18/09/2024) and Institutional ethics committee, Mahatma Gandhi Institute of Medical Sciences, Sevagram, Wardha (MGIMS/IEC/COMMED/187/2024 dated 31/8/2024) before the initiation of the study.

## Results

### Contact tracing and enlisting household contacts

A total of 2,23,827 TB patients were notified in 2023 in Maharashtra. After excluding people with extra pulmonary TB (n=80656), those with disease site not reported (n=9944), those with enrolment dates outside 2023 (n=32) and outliers with respect to number of HHC (n=28), we included 1,33,167 PwPTB for analysis. Contact tracing was done for 1,12,034 (84%) PwPTB through which a total of 4,06,291 HHC were identified. Of the identified HHC, 3,81,326 (94%) were aged above 6 years. The median number of enlisted HHC per PwPTB was 3 (IQR: 2-4).

### Tuberculosis preventive treatment care cascade

Among the 1,85,502 HHC eligible for TPT, 1,01,325 (55%) were put on treatment. Overall, 4,06,291 HHC were enlisted through contact tracing and of them 3,62,809 (95%) were screened for symptoms of TB (Figure 1). Among the screened contacts, 7632 (2%) were symptomatic, of which 6348 (83%) were evaluated for presence of TB disease. Of the symptomatic contacts who were evaluated for presence of TB disease, 799 (13%) were diagnosed with TB, of whom 649 (81%) were initiated on TB treatment. Of the total HHC, 1,85,502 (46%) were eligible for TPT among whom, 1,01,325 (55%) were initiated on TPT. This amounts to around 25% of the enlisted HHC. Of those who were started on TPT, 89,506 (88%) were initiated on the same day of the diagnosis of the index PwPTB.

**Figure 1:**
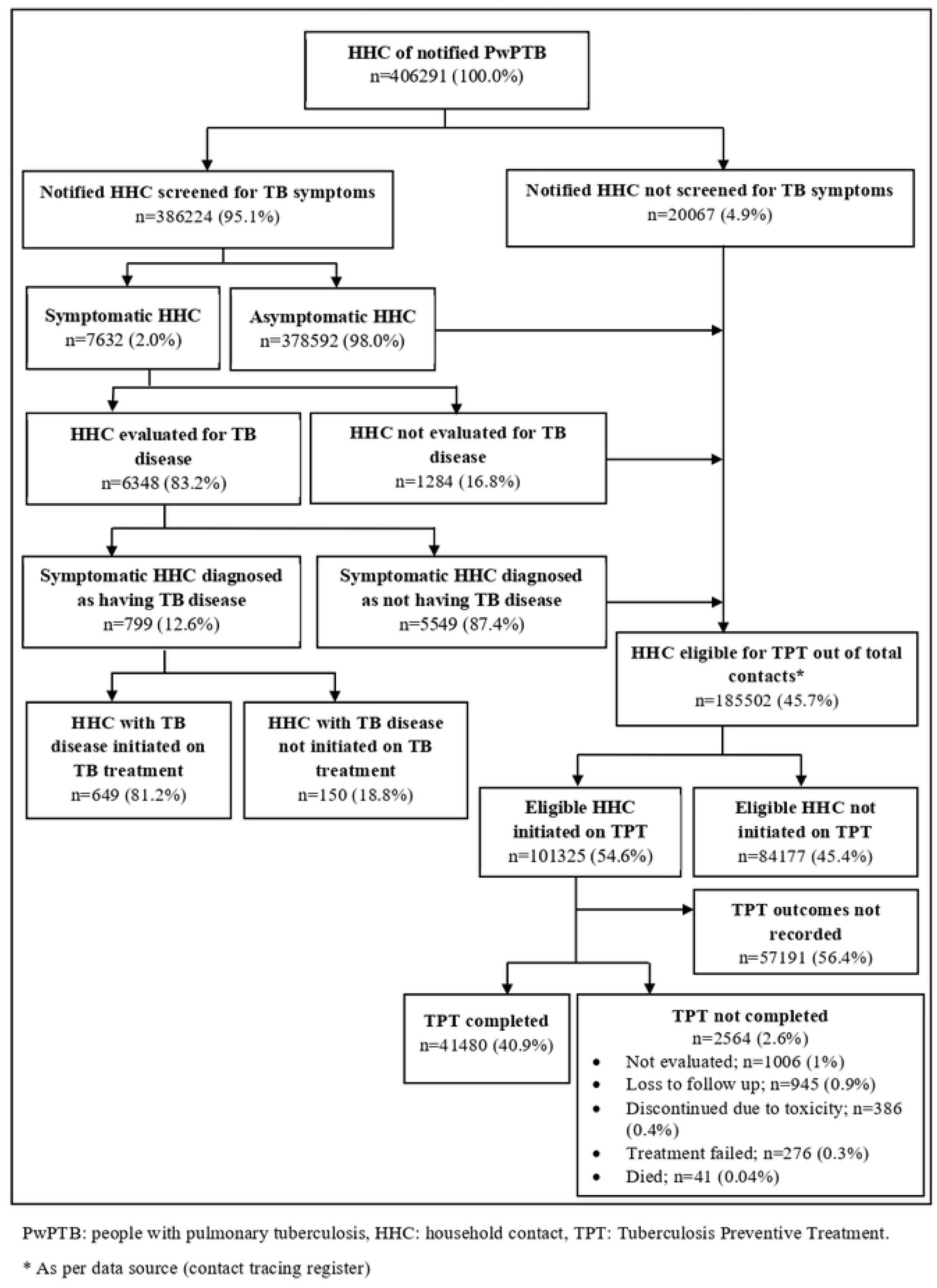
Cascade of care for Tuberculosis preventive treatment among household contacts of PwPTB in Maharashtra, India during the year 2023 (N=4,06,291)

Treatment outcomes were reported for 44,134 (44%) HHC. While 41,480 (41%) contacts successfully completed the TPT treatment, 2,564 (3%) contacts had not completed TPT. The characteristics of the HHC initiated on TPT and respective treatment outcomes are described in table 1. Six TPT regimens were administered with majority receiving 6H regimen (86497; 85%) followed by 3HP regimen (5364; 5%). TPT completion was similar for all age groups and for both the genders. It was higher among those on shorter regimens i.e. 3HP (78%), 4R (72%), and 3HR (56%) as compared to those on 6H regimen (41%). Treatment outcomes for 56% of the contacts was not recorded. Missing data was higher if the index patient sought care from private sector (63%), among contacts of clinically diagnosed cases (70%), and among those administered 6H regimen (57%) as compared to the other shorter regimens.

**Table 1:**
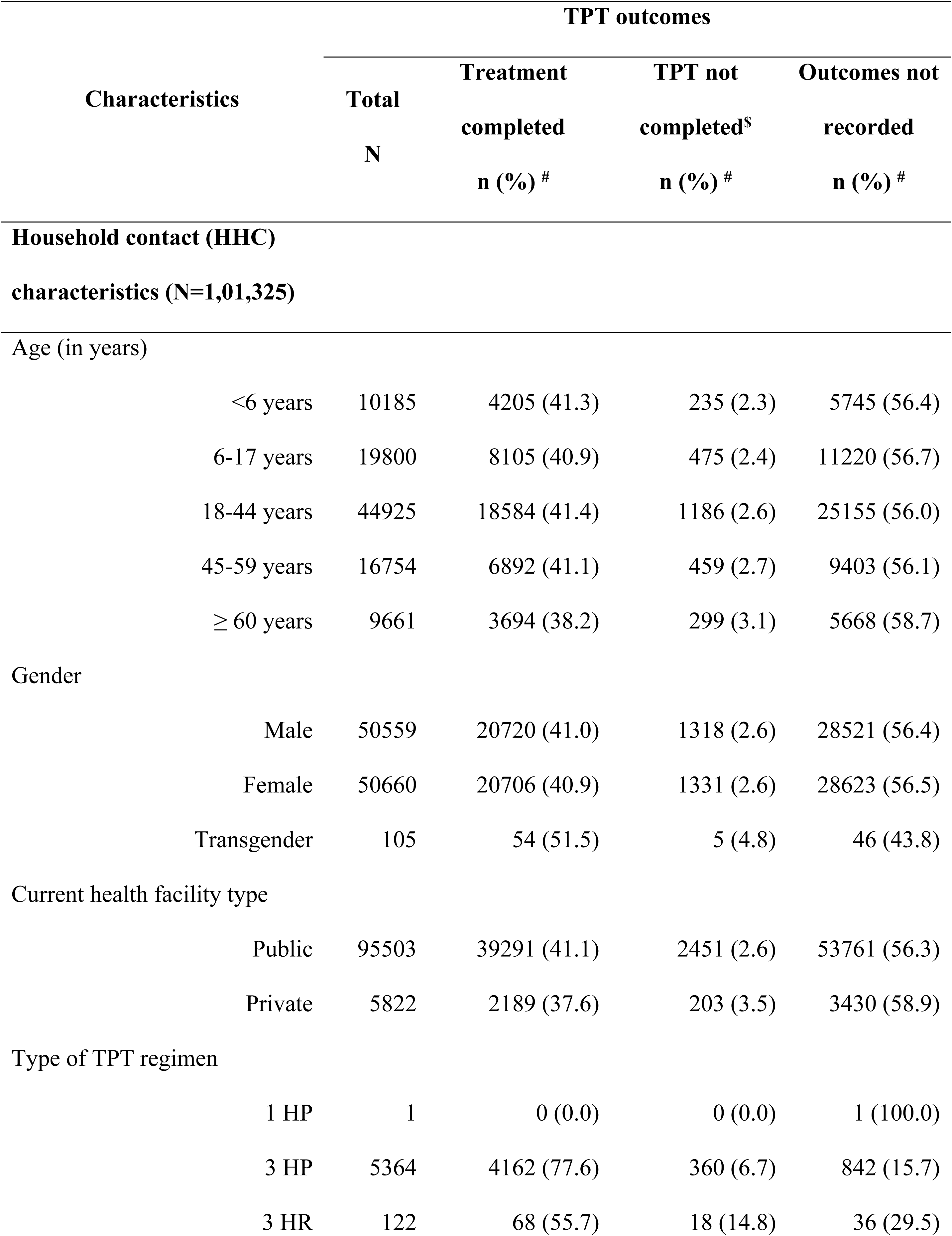
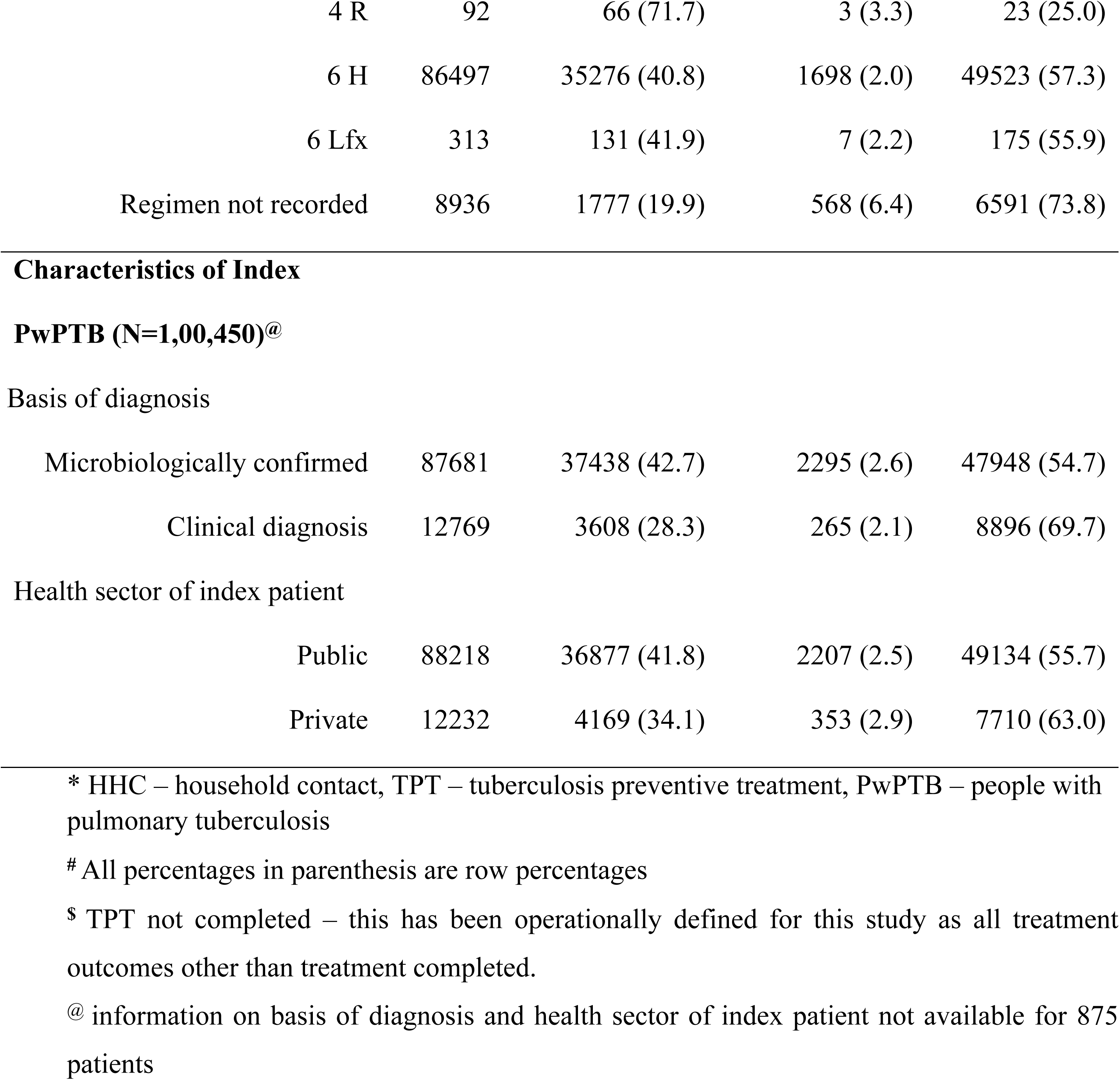
Select characteristics of HHC of PwPTB by TPT treatment outcomes in Maharashtra, India during the year 2023.

| Characteristics | TPT outcomes |  |  |  |
| --- | --- | --- | --- | --- |
|  | Total | Treatment | TPT not | Outcomes not |
|  | N | completed | completed<sup>s</sup> | recorded |
|  |  | n (%) # | n (%) # | n (%) # |
| **Household contact (HHC)** |  |  |  |  |
| **characteristics (N=1,01,325)** |  |  |  |  |
| Age (in years) |  |  |  |  |
| <6 years | 10185 | 4205 (41.3) | 235 (2.3) | 5745 (56.4) |
| 6-17 years | 19800 | 8105 (40.9) | 475 (2.4) | 11220 (56.7) |
| 18-44 years | 44925 | 18584 (41.4) | 1186 (2.6) | 25155 (56.0) |
| 45-59 years | 16754 | 6892 (41.1) | 459 (2.7) | 9403 (56.1) |
| ≥ 60 years | 9661 | 3694 (38.2) | 299 (3.1) | 5668 (58.7) |
| Gender |  |  |  |  |
| Male | 50559 | 20720 (41.0) | 1318 (2.6) | 28521 (56.4) |
| Female | 50660 | 20706 (40.9) | 1331 (2.6) | 28623 (56.5) |
| Transgender | 105 | 54 (51.5) | 5 (4.8) | 46 (43.8) |
| Current health facility type |  |  |  |  |
| Public | 95503 | 39291 (41.1) | 2451 (2.6) | 53761 (56.3) |
| Private | 5822 | 2189 (37.6) | 203 (3.5) | 3430 (58.9) |
| Type of TPT regimen |  |  |  |  |
| 1 HP | 1 | 0 (0.0) | 0 (0.0) | 1 (100.0) |
| 3 HP | 5364 | 4162 (77.6) | 360 (6.7) | 842 (15.7) |
| 3 HR | 122 | 68 (55.7) | 18 (14.8) | 36 (29.5) |
| 4 R | 92 | 66 (71.7) | 3 (3.3) | 23 (25.0) |
| 6 H | 86497 | 35276 (40.8) | 1698 (2.0) | 49523 (57.3) |
| 6 Lfx | 313 | 131 (41.9) | 7 (2.2) | 175 (55.9) |
| Regimen not recorded | 8936 | 1777 (19.9) | 568 (6.4) | 6591 (73.8) |
| Basis of diagnosis |  |  |  |  |
| Microbiologically confirmed | 87681 | 37438 (42.7) | 2295 (2.6) | 47948 (54.7) |
| Clinical diagnosis | 12769 | 3608 (28.3) | 265 (2.1) | 8896 (69.7) |
| Health sector of index patient |  |  |  |  |
| Public | 88218 | 36877 (41.8) | 2207 (2.5) | 49134 (55.7) |
| Private | 12232 | 4169 (34.1) | 353 (2.9) | 7710 (63.0) |

\* HHC – household contact, TPT – tuberculosis preventive treatment, PwPTB – people with pulmonary tuberculosis

## Discussion

There were considerable losses in the TPT care cascade in Maharashtra, including contact tracing of index PwPTB and TPT completion of HHC. Around half of the eligible contacts were initiated on TPT. TPT outcomes for over half of the HHC put on treatment were not recorded in the *Ni-kshay* portal. The proportion of missing treatment outcomes was higher for those initiated on 6H regimen, private sector patients, and clinically diagnosed cases.

In our study, TPT was not initiated in about half of the eligible HHC. Overall, one of every four enlisted HHC received TPT in our settings. This is similar to the national status of HHC of bacteriologically confirmed PwPTB receiving TPT (24%) as per India TB report 2023[17] and slightly better than the global proportion (21%).[21] Alsdurf et al.[22] in their meta-analysis of 70 cohorts of people intended for TBI evaluation and treatment found that a small proportion of eligible persons (5%) do not receive TPT. Fear of side effects[14,16], low risk perception among the HHC, and lack of proper information and counselling[12,23] are some of the important reasons for low acceptance of TPT. High workload of health care staff may also be one of the reasons why programmes fail to initiate HHC on TPT.[24]

Contact tracing was not done for all the index PwPTB. The state enlists a median 3 contacts per PwPTB. The mean contacts enlisted per PwPTB from other Indian studies was 3.8 from Maharashtra[14], 3.1 from Karnataka[15], 2.9 from Delhi[16]. Contact tracing and enumeration of HHC is the first and most critical step of the cascade. National programmes across countries often tend to underreport the number of contacts at risk.[24] High workload[16], unavailability of family members during home visit[14], and stigma are some of the reasons of loss at this step. Evaluation of household contacts for TB disease or TBI is the next step in the cascade. The effectiveness of TPT is dependent on the identification of people who are eligible and would benefit most from it.[24] Alsdurf et al. [22] found that only around two thirds of those intended for evaluation/screening actually undergo the same. Limited availability of TBI testing services, unavailability of identified contacts for tests, lack of awareness, limited involvement of routine healthcare staff,[14] and stigma[15] are some reasons of losses at this stage.

In our study the reported TPT completion was low. Missing data about treatment outcomes, the pattern of which we are not sure about, limits our ability to comment on over or under reporting of non-completion of TPT. The TPT outcomes were better for shorter regimens. Systematic reviews have demonstrated better TPT completion with shorter regimens.[25,26] Treatment related serious adverse events have been similar or lesser in shorter regimen as compared to 6H regimen[25–27]. Studies from India[16] and Peru[28], suggest that beneficiaries preferred shorter regimen as the dosage schedule are easy to remember and less disruptive.

For over half the contacts initiated on TPT, the outcomes were not recorded/ updated. TB programme workforce is accustomed to monitor treatment outcomes among individuals with disease and hence, monitoring healthy contacts for treatment, as in the case of TPT may be challenging and a possible reason for missing outcomes.[24] In our study, the recording of outcomes among HHC of clinically diagnosed cases was lesser than that among HHC of microbiologically confirmed ones and TPT completion was also lesser. The risk of TPT non-completion among HHC of clinically diagnosed PwPTB was 1.6 times as compared to HHC of microbiologically confirmed PwPTB.[11] The national guidelines recommend initiation of TPT among HHC of all, however, the India TB report does not show TPT indicators for HHC of clinically confirmed PwPTB.[17] The India TB report needs to be aligned to the national guidelines to minimize any complacency that may result due to such differences. The recording of outcomes was lower among patients and/or contacts seeking care from private sector. Such contacts are also at higher risk of TPT non-completion and need better monitoring.[11]

Our study had few limitations. First, we were unable to analyze the delays in the cascade and time to events due to missing outcomes and their dates. Second, the data from contact tracing register was an aggregate one whereas from the other two registers was individual level, due to which, we were unable to understand the proportions of losses from the cascade following symptomatic screening for TB and during TBI evaluation. Lastly, though TPT is also being given to people living with HIV, we did not include them in the analysis.

The aspect of TPT provision for HHC > 6 years, is a newer addition in the programme. Further streamlining and strengthening this aspect will require training of the routine health care staff including the private sector for contact tracing, TBI evaluation and follow ups. Decentralization and better involvement of health and wellness centres for TPT is needed for its effectively implementation. Supportive supervision and monitoring also needs to be strengthened for TPT. Since the quantum of TPT beneficiaries is large, involvement of health and wellness centres and frontline workers in data recording through a simpler version of electronic data capturing tools may prove beneficial. This will not only promote real time data capturing, but has the potential to be used as a monitoring as well as decision making tool. Lastly, the programme should consider including TPT indicators for HHC of all PwPTB (microbiologically confirmed and clinically diagnosed) instead of only microbiologically confirmed PwPTB in the annual TB report.

## Conclusion

From this statewide data analysis, we identified losses in the TPT care cascade and incomplete recording of information, particularly treatment outcomes. Capacity building of the health care staff responsible for implementing TPT, routine review of TPT indicators and data completion, strengthening the monitoring and supportive supervision and further improvement in the way data is captured in the web-based data management system is required. We recommend that annual India TB report should include TPT related information for HHC of all PwPTB, thereby aligning the indicators in annual reports with the guidelines. We also propose to conduct an implementation study to record selective information by frontline workers/ health and wellness centers utilizing real time electronic data capturing mechanisms and assess the delays and treatment outcomes in the TPT care cascade and in the process strengthen its implementation as well.

## Data Availability

The data that support the findings of this study have been deposited in figshare at 10.6084/m9.figshare.28785116 (notification and contact tracing register) and 10.6084/m9.figshare.28785119 (TPT register)

https://10.6084/m9.figshare.28785119

https://10.6084/m9.figshare.28785116

## Acknowledgements

We would like to thank Dr Ashok Randive and Mr Waghmare from the state NTEP office, Mr Sumant Dhoble and Mr Punvatkar from District TB centre, Wardha for their help in data acquisition and inputs during the process of data cleaning and analysis.

This operational/ implementation research that resulted in this manuscript was conducted through the Structured Operational Research and Training Initiative (SORT IT), a global partnership led by the Special Program for Research and Training in Tropical Diseases at the World Health Organization (WHO/TDR). The model is based on a course developed jointly by the International Union Against Tuberculosis and Lung Disease (The Union) and Medécins sans Frontières (MSF/Doctors Without Borders). This specific SORT IT course which resulted in this publication was part of year one of ICMR-National Institute of Epidemiology (ICMR-NIE) led TB SORT IT course 2024-26, with support and guidance from India’s Central TB Division and WHO India. It was jointly developed and implemented by: ICMR-National Institute of Epidemiology (ICMR-NIE), Chennai, India; ICMR-National Institute for Research in Tuberculosis (ICMR-NIRT), Chennai, India; Post Graduate Institute of Medical Education and Research (PGIMER), Chandigarh, India; FIND, New Delhi, India; Baroda Medical College, Vadodara, India; Narotam Sekhsaria Foundation, Mumbai, India; Government Medical College, Shahdol, India; All India Institute of Medical Sciences (AIIMS), Madurai, India; All India Institute of Medical Sciences (AIIMS), Bhathinda, India; Yenepoya Medical College, Mangaluru, India; and GMERS Gotri Medical College, Vadodara, India.

